# Development and external validation of a diagnostic model for in-hospital bleeding in patients with acute ST elevation myocardial infarction

**DOI:** 10.1101/2020.05.28.20115501

**Authors:** Yong Li

**Affiliations:** Emergency and Critical Care Center, Beijing Anzhen Hospital, Capital Medical University, Beijing 100029, China

**Keywords:** coronary disease, ST elevation myocardial infarction, hemorrhage, nomogram

## Abstract

**Background:** Bleeding complications in patients with acute ST segment elevation myocardial infarction (STEMI) are associated with an increased risk of subsequent adverse consequences. We want to develop and externally validate a diagnostic model of in-hospital bleeding in the population of unselected real-world patients with acute STEMI.

**Methods:** Design: Multivariable logistic regression of a cohort for hospitalized patients with acute STEMI. Setting: Emergency department ward of a university hospital. Participants: Diagnostic model development: Totally 4262 hospitalized patients with acute STEMI from January 2002 to December 2013 in Beijing Anzhen Hospital, Capital Medical University. External validation: Totally 6015 hospitalized patients with acute STEMI from January 2014 to August 2019 in Beijing Anzhen Hospital, Capital Medical University. Outcomes: All-cause in-hospital bleeding not related to coronary artery bypass graft surgery or catheterization.

**Results:** In-hospital bleeding occurred in 2.6% (112/4262) of patients in the development data set (117/6015) of patients in the validation data set. The strongest predictors of in-hospital bleeding were advanced age and high Killip classification. We developed a diagnostic model of in-hospital bleeding. The area under the receiver operating characteristic ROC curve (AUC) was 0.777±0.021, 95% confidence interval(CI) = 0.73576 ~ 0.81823. We constructed a nomograms using the development database based on age, and Killip classification. The AUC was 0.7234±0.0252,

95% CI = 0.67392 ~ 0.77289 in the validation data set. Discrimination, calibration, and decision curve analysis were satisfactory.

**Conclusions:** We developed and externally validated a moderate diagnostic model of in-hospital bleeding in patients with acute STEMI.

We registered this study with WHO International Clinical Trials Registry Platform (ICTRP) (registration number: ChiCTR1900027578; registered date: 19 Novmober 2019). http://www.chictr.org.cn/edit.aspx?pid=45926&htm=4.

## Background

Hemorrhagic complications occurred in nearly 8.5% of patients with acute ST segment elevation myocardial infarction (STEMI) during hospitalization.^[1, 2]^ Bleeding events are associated with an increased risk of adverse outcomes in patients with STEMI.^[3–6]^ Prevention of bleeding may represent an achievable step. We want to develop and externally validate a diagnostic model of in-hospital bleeding in the population of unselected real-world patients with acute STEMI. The aim of our study was 4-fold: (1) to identify predictive factors; (2) to develop a diagnostic model; (3) to create a nomogram and (4) to externally validate diagnostic model.

## Methods

We used a Type 2b predictive model study, which covered by Transparent Reporting of a multivariable prediction model for Individual Prognosis Or Diagnosis (TRIPOD) statement.^[7]^ The data was divided into two groups non-randomly according to time: one group was used to develop a prediction model, and the other group was used to evaluate its prediction performance.^[7]^Type 2b was called”external verification study”.^[7]^

The derivation cohort was 4262 hospitalized patients with acute STEMI from January 2002 to December 2013 in Beijing Anzhen Hospital, Capital Medical University.

The validation cohort was 6015 hospitalized patients with acute STEMI from January 2014 to August 2019 in Beijing Anzhen Hospital, Capital Medical University.

Inclusion criteria: 1. hospitalized patients with acute STEMI; 2. age of more than 18 years. We established the diagnosis of acute myocardial infarction (AMI) and STEMI base on the fourth universal definition of myocardial infarction.^[8]^

Exclusion criteria: none.

It was a retrospective analysis and informed consent was waived by Ethics Committee of Beijing Anzhen Hospital Capital Medical University.

Outcome of interest was all-cause in-hospital bleeding not related to coronary artery bypass graft surgery or catheterization during hospitalization, as defined according to the Bleeding Academic Research Consortium criteria2, 3, and 5.^[4]^ The presence or absence of in-hospital bleeding was decided blinded to the predictor variables and based on the medical record.

We selected 13 predictor according to clinical relevance and the results of baseline descriptive statistics. The potential candidate variables were age, sex, Killip classification, atrioventricular block (AVB), atrial fibrillation(AF), underwent percutaneous coronary intervention(PCI)during hospitalization, and medical history such as hypertension, diabetes, myocardial infarction, PCI, coronary artery bypass grafting (CABG), cerebrovascular disease (HCD), and chronic kidney disease(CKD). All of them based on the medical record. AF defined as all type of AF during hospitalization. AVB defined as all type of AVB during hospitalization.

Some people suggest that each candidate variable has at least 10 events for model derivation and at least 100 events for validation studies.^[7]^Our number of samples and events exceeds all approaches used to determine sample size, so it is expected to provide a very reliable estimate. In order to ensure the reliability of the data, we excluded patients who lacked information on key predictors: age and Kilip classification. The reason for excluding all patients was the lack of Killip classification.

### Statistical analysis

We kept all continuous data as continuous and retained on the original scale. Based on the variables significantly generated by univariate logistic regression, we constructed a multivariate logistic regression model using the backward variable selection method.We used the Akanke information criterion (AIC) and Bayesian information criterion(BIC)to select predictors. It considers model fitting and penalizes the estimated number of parameters, which is equivalent to using α = 0.157.^[7]^ We assessed the predictive performance of the diagnostic model in the validation data sets by examining measures of discrimination, calibration, and decision curve analysis (DCA).^[7, 9]^ Discrimination is the ability of the diagnostic model to differentiate between patients who with and without in-hospital bleeding. This measure was quantified by calculating the area under the receiver operating characteristic (ROC) curve (AUC).^[7]^Calibration refers to how closely the predicted in-hospital bleeding agrees with the observed in-hospital bleeding.^[7]^ The Brier score was an aggregate measure of disagreement between the observed outcome and a prediction— the average squared error difference. We used DCA to describe and compare the clinical effects of diagnostic model.^[7]^We performed statistical analyses with STATA version 15.1 (StataCorp, College Station, TX), R version 4.0.0(R Development Core Team; http://www.r-project.org) and the RMS package developed by Harrell (Harrell et al). All tests were two-sided and a P value <0.05 was considered statistically significant.

## Results

We drew a flow diagrams (Figure 1).

**Figure 1.**
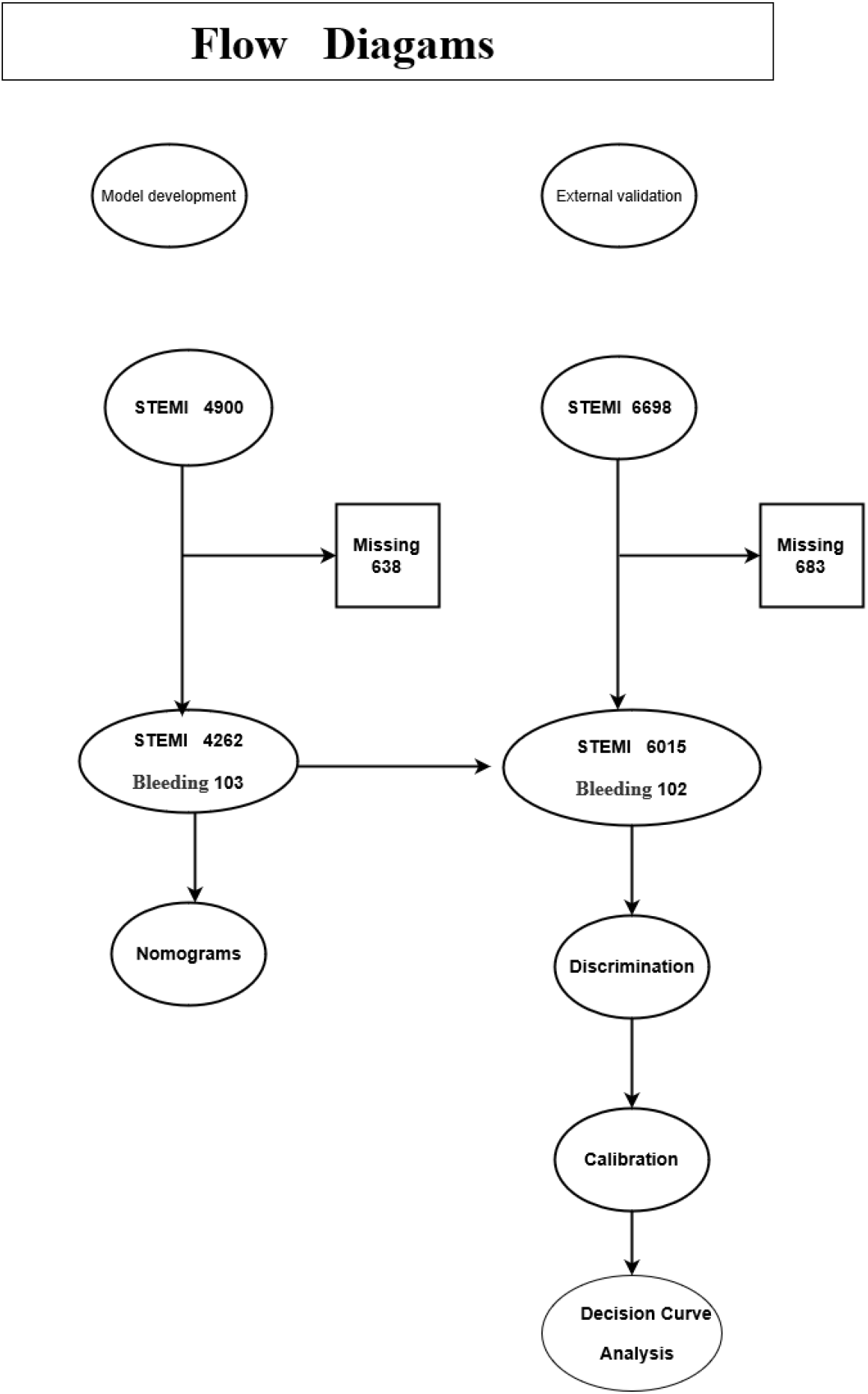
Flow diagrams.

In the development data set, a total of 2.6% (112/4262) of hospitalized patients experienced in-hospital bleeding. The patient’s baseline characteristics were shown in Table 1. Nine variables (age, sex, Killip classification, AVB, AF, history of CABG, history of diabetes, history of CKD, and underwent PCI during hospitalization)were significant differences in the two groups of patients (p < 0. 157). After application of backward variable selection method, AIC, and BIC, age remained as a significant independent predictors of in-hospital bleeding; Killip classification remained as a rank variable of in-hospital bleeding. Results were shown in Table 2 and Table 3.

**Table 1.**
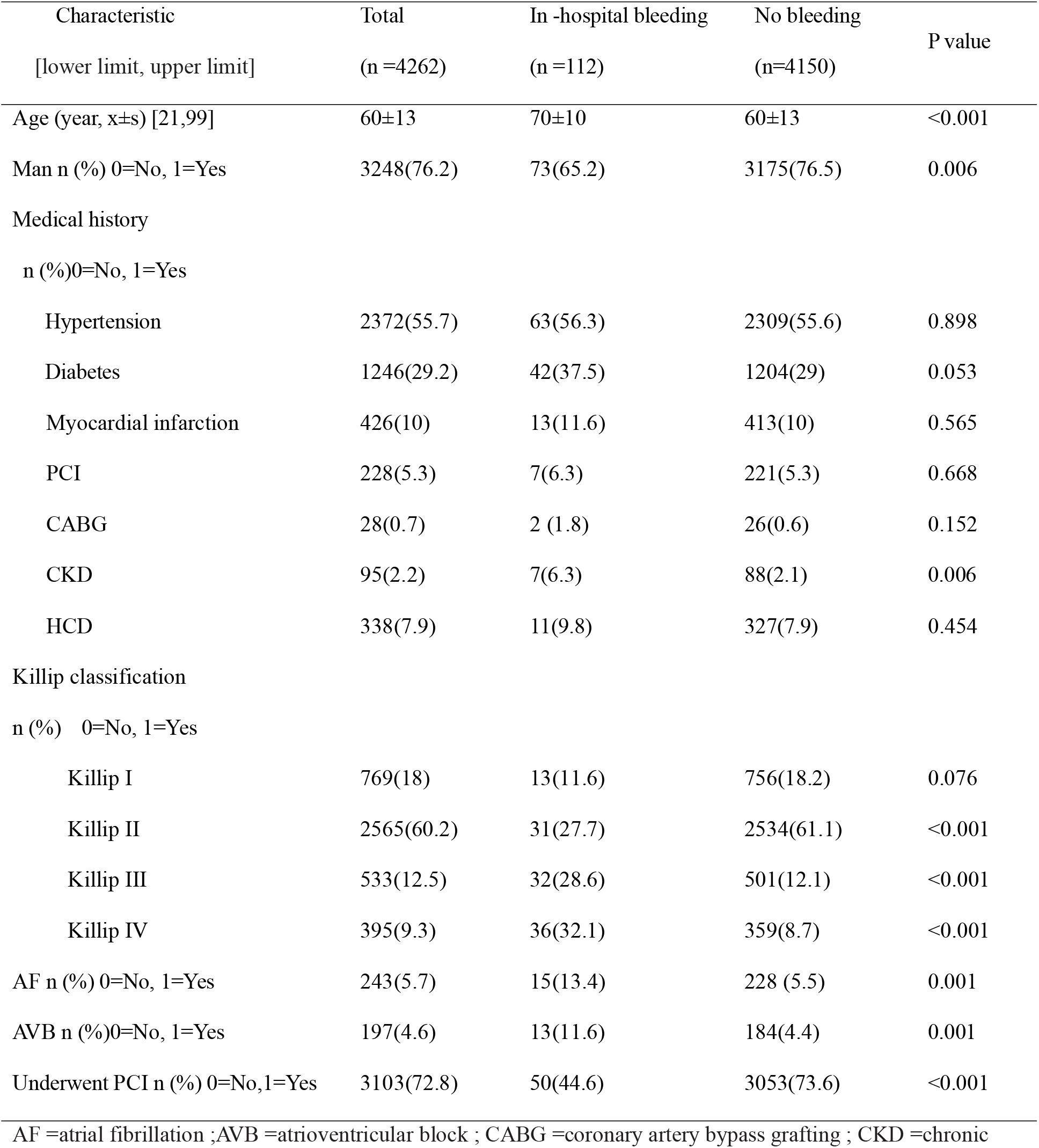
Demographic and clinical characteristics of patients with and without in-hospital bleeding in the development data sets.

| Characteristic<br>[lower limit, upper limit] | Total<br>(n =4262) | In -hospital bleeding<br>(n =112) | No bleeding<br>(n=4150) | P value |
| --- | --- | --- | --- | --- |
| Age (year, x±s) [21,99] | 60±13 | 70±10 | 60±13 | <0.001 |
| Man n (%) 0=No, 1=Yes | 3248(76.2) | 73(65.2) | 3175(76.5) | 0.006 |
| Medical history |  |  |  |  |
| n (%)0=No, 1=Yes |  |  |  |  |
| Hypertension | 2372(55.7) | 63(56.3) | 2309(55.6) | 0.898 |
| Diabetes | 1246(29.2) | 42(37.5) | 1204(29) | 0.053 |
| Myocardial infarction | 426(10) | 13(11.6) | 413(10) | 0.565 |
| PCI | 228(5.3) | 7(6.3) | 221(5.3) | 0.668 |
| CABG | 28(0.7) | 2 (1.8) | 26(0.6) | 0.152 |
| CKD | 95(2.2) | 7(6.3) | 88(2.1) | 0.006 |
| HCD | 338(7.9) | 11(9.8) | 327(7.9) | 0.454 |
| Killip classification |  |  |  |  |
| n (%) 0=No, 1=Yes |  |  |  |  |
| Killip I | 769(18) | 13(11.6) | 756(18.2) | 0.076 |
| Killip II | 2565(60.2) | 31(27.7) | 2534(61.1) | <0.001 |
| Killip III | 533(12.5) | 32(28.6) | 501(12.1) | <0.001 |
| Killip IV | 395(9.3) | 36(32.1) | 359(8.7) | <0.001 |
| AF n (%) 0=No, 1=Yes | 243(5.7) | 15(13.4) | 228 (5.5) | 0.001 |
| AVB n (%)0=No, 1=Yes | 197(4.6) | 13(11.6) | 184(4.4) | 0.001 |
| Underwent PCI n (%) 0=No,1=Yes | 3103(72.8) | 50(44.6) | 3053(73.6) | <0.001 |

**Table 2.** Predictor of in-hospital bleeding obtained from multivariable logistic regression models (odds ratio) in the development data set.

| In -hospital bleeding | Odds ratio | Std.Err | Z | P>\| Z \| | 95% CI |
| --- | --- | --- | --- | --- | --- |
| Age | 1.047443 | .0095986 | 5.06 | <0.001 | 1.028798~1.066426 |
| Killip III | 3.265072 | .8100203 | 4.77 | <0.001 | 2.007804~5.309632 |
| Killip IV | 5.132613 | 1.240357 | 6.77 | <0.001 | 3.196212~8.242169 |
| _cons | .0007621 | .0004685 | -11.68 | <0.001 | .0002285 ~.0025424 |

CI =confidence interval.

**Table 3.** Predictor of in-hospital bleeding obtained from multivariable logistic regression models (Coef) in the development data sets.

| In -hospital bleeding | Coef | Std.Err | Z | P>\| Z \| | 95% CI |
| --- | --- | --- | --- | --- | --- |
| Age | .0463523 | .0091638 | 5.06 | <0.001 | .0283915~ .0643131 |
| Killip III | 1.183282 | .2480865 | 4.77 | <0.001 | .6970414~1.669523 |
| Killip IV | 1.635615 | .2416619 | 6.77 | <0.001 | 1.161966 ~ 2.109263 |
| _cons | -7.179377 | .6146614 | -11.68 | <0.001 | -8.384092~-5.974663 |

CI =confidence interval.

According to the above risk factors, we can calculate the predicted probability of in-hospital bleeding using the following formula: P = 1/(1+exp(−(−7.179377 +.0463523*AGE(year) +1.183282* KIII+ 1.635615* KIV))). KIII = Killip III(0 = No, 1 = Yes), KIV = Killip IV (0 = No, 1 = Yes). The ROC curve was drawn (Figure 2).AUC was 0.777±0.021, 95% confidence interval(CI) = 0.73576 ~ 0.81823.

**Figure 2.**
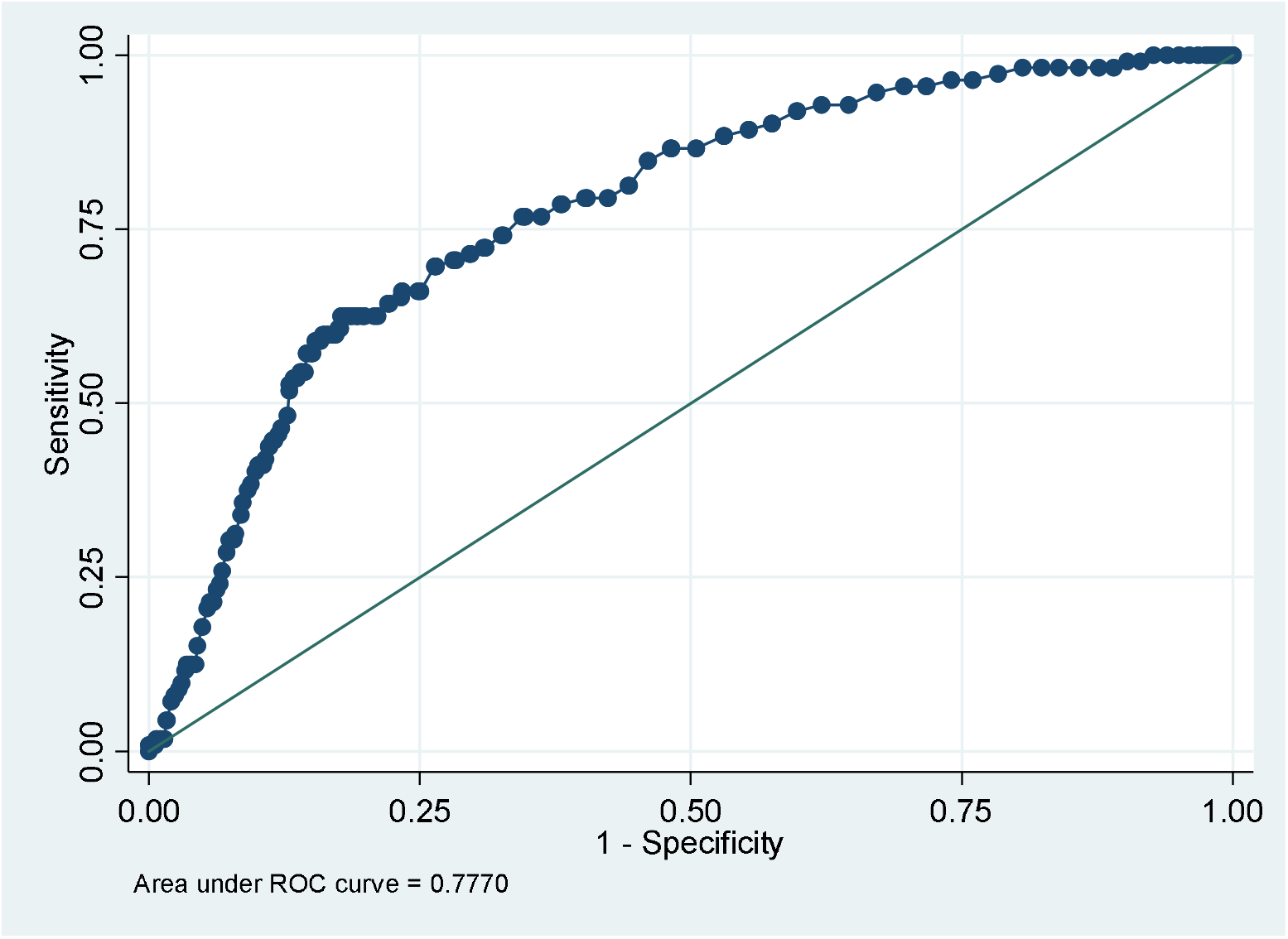
Receiver-operating characteristics curve in identifying patients with in-hospital bleeding in the development dataset.

We constructed the nomogram (Figure 3) using the development database based on a independent prognostic marker and a rank variable: age and Killip classification.

**Figure 3.**
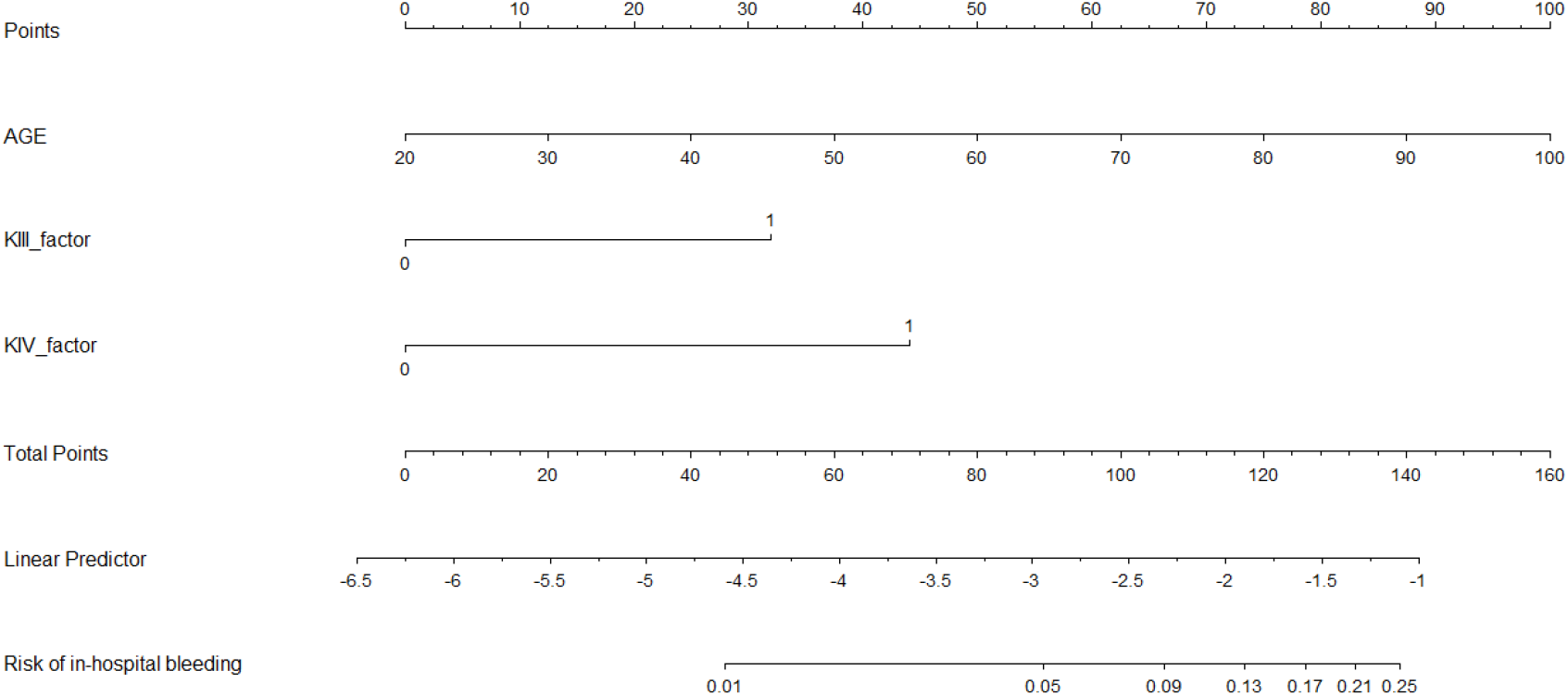
A nomograms for predicting in-hospital bleeding in patients with acute STEMI AGE = Age(year); KIII-factor = Killip III; KIV-factor = Killip IV.

Totally 1.9% (117/6015) hospitalized patients suffered in-hospital bleeding in the validation data sets. Baseline characteristics of the patients were shown in Table 4.We can calculate the predicted probability of in-hospital bleeding using the following formula: P = 1/(1+exp(−(−7.179377 +.0463523*AGE(year) +1.183282* KIII+ 1.635615* KIV))). KIII = Killip III(0 = No, 1 = Yes), KIV = Killip IV (0 = No, 1 = Yes). We drew the ROC curve (Figure 4). AUC was 0.7234±0.0252, 95% CI = 0.67392 ~ 0.77289.

**Table 4.** Demographic and clinical characteristics of patients with and without in-hospital bleeding in the validation data sets.

| Characteristic<br>[lower limit, upper limit] | Total<br>(n =6015) | In-hospital bleeding<br>(n =117) | No bleeding<br>(n=5898) | P value |
| --- | --- | --- | --- | --- |
| Age (year, x±s) [21,92] | 59±12 | 64±12 | 58±12 | <0.001 |
| Man n (%) 0=No, 1=Yes | 4894(81.4) | 86(73.5) | 4808(81.5) | 0.029 |
| Medical history, n (%)<br>0=No, 1=Yes |  |  |  |  |
| Hypertension | 3427(57) | 65(55.6) | 3362(57) | 0.754 |
| Diabetes | 1822(30.3) | 40(34.2) | 1782(30.2) | 0.355 |
| Myocardial infarction | 433(7.2) | 14(12) | 419(7.1) | 0.047 |
| PCI | 575(9.6) | 18(15.4) | 557(9.4) | 0.033 |
| CABG | 51(0.8) | 3 (2.6) | 48(0.8) | 0.053 |
| CKD | 145(2.4) | 4(3.4) | 141(2.4) | 0.475 |
| HCD | 421(7) | 12(10.3) | 409(6.9) | 0.166 |
| Killip classification<br>n (%) 0=No, 1=Yes |  |  |  |  |
| Killip I | 4234(70.4) | 45(38.5) | 4189(71) | <0.001 |
| Killip II | 1188(19.7) | 31(26.5) | 1157(19.6) | 0.066 |
| Killip III | 266(4.4) | 11(9.4) | 255(4.3) | 0.010 |
| Killip IV | 330(5.5) | 30(25.6) | 300(5.1) | <0.001 |
| AF n (%) 0=No, 1=Yes | 275(4.6) | 12(10.3) | 263 (4.5) | 0.004 |
| AVB n (%) 0=No, 1=Yes | 119(2) | 3(2.6) | 116(2) | 0.647 |
| Underwent PCI n (%) 0=No, 1=Yes | 4564(75.9) | 70(59.8) | 4494(76.2) | <0.001 |

Abbreviations as in Table 1.

**Figure 4.**
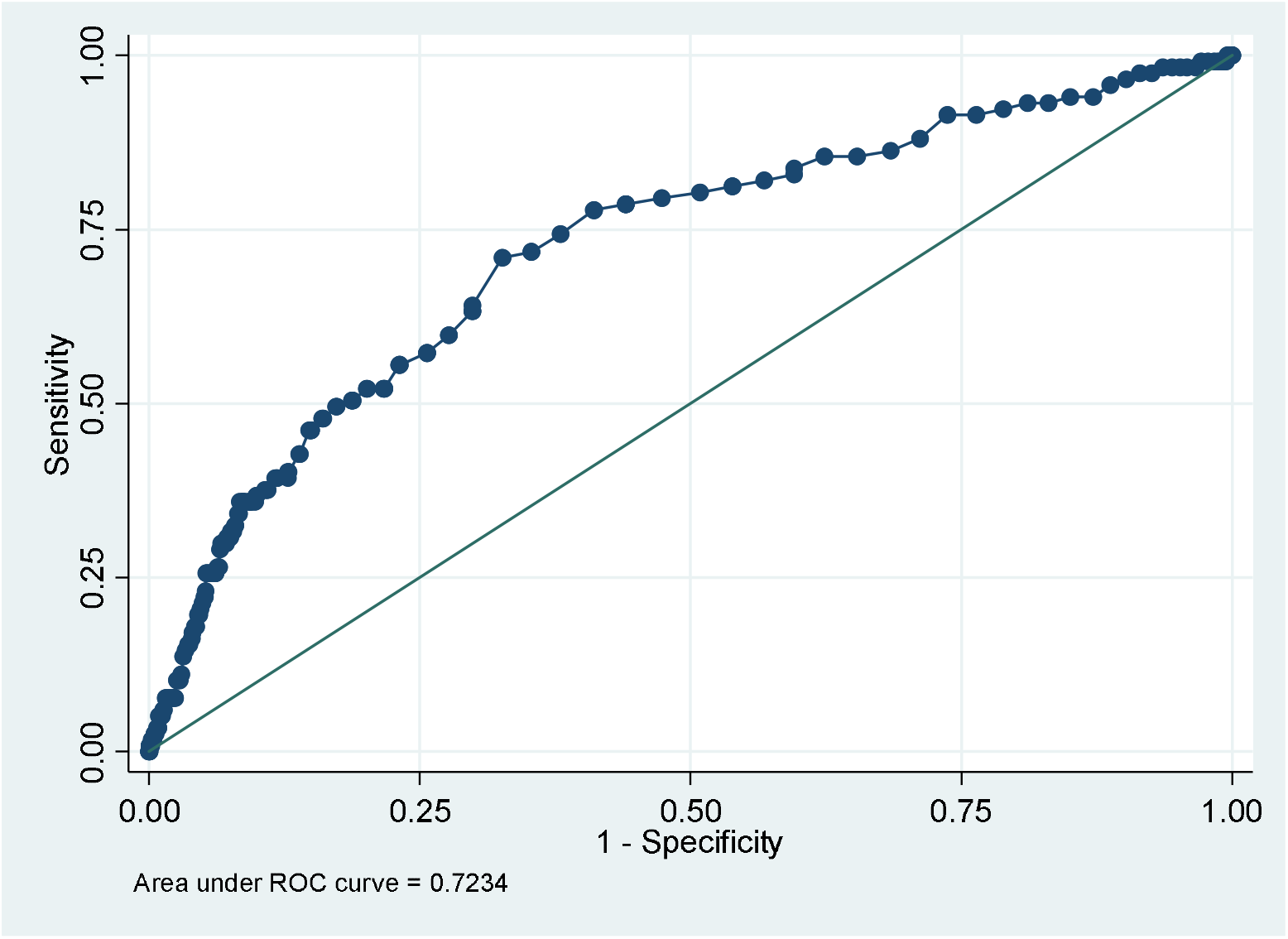
Receiver-operating characteristics curve in identifying patients with in-hospital bleeding in the validation data sets.

We drew a calibration plot (Figure 5) with distribution of the predicted probabilities for individuals with and without in-hospital bleeding in the validation data sets.Hosmer-Lemeshow chi2(10) = 10.64,Prob > chi2 = 0. 3859 >0.05. Brier score = 0.0188<0.25.

**Figure 5.**
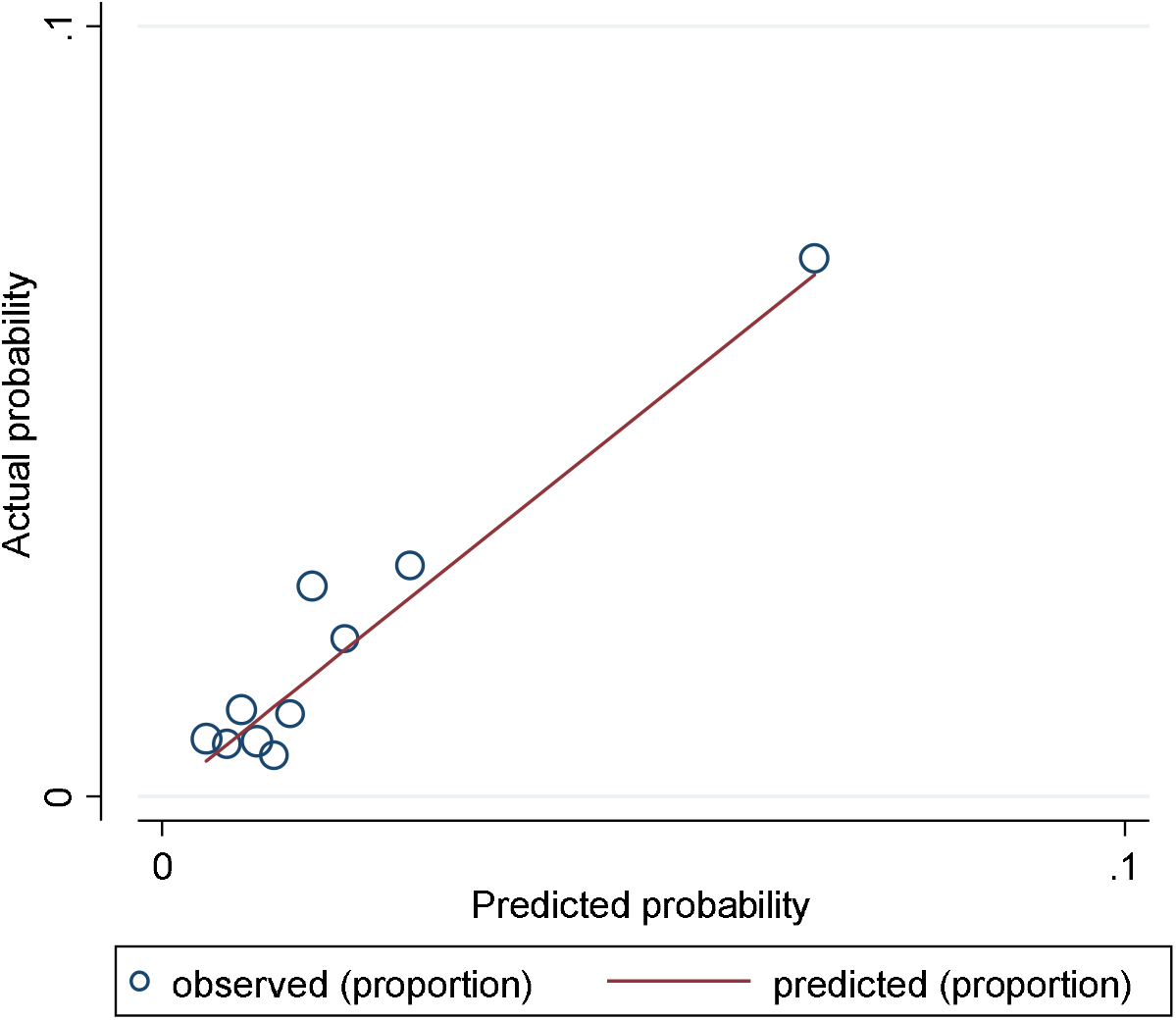
A calibration plot with distribution of the predicted probabilities for individuals with and without in-hospital bleeding in the validation data sets.

DCA(Figure 6) in the validation data sets.

**Figure 6.**
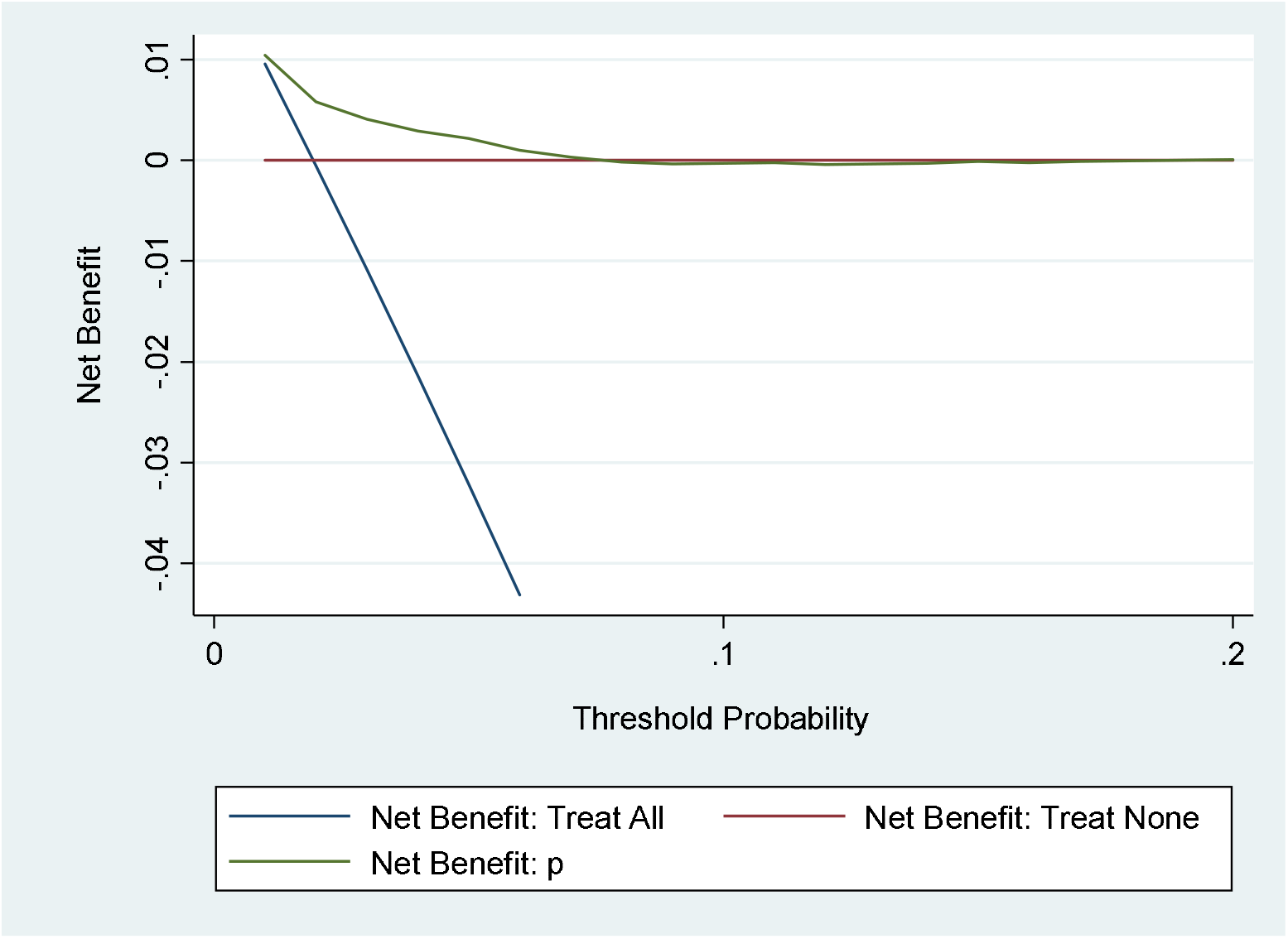
DCA in the validation data sets.

## Discussion

We assessed the predictive performance of the diagnostic model in the validation data sets by examining measures of discrimination, calibration, and DCA. AUC was 0.7234±0.0252, 95% CI = 0.67392 ~ 0.77289 in the validation data sets. Hosmer-Lemeshow chi2(10) = 10.64, Prob > chi2 = 0. 3859 >0.05. Brier score <0.25. Discrimination, calibration, and DCA were satisfactory. In our study, advanced age and high Killip classification are associated with an increased risk of in-hospital bleeding in patients with acute STEMI. We can use the formula or nomogram to predicte in-hospital bleeding. We can use specific strategies to reduce the risk of in-hospital bleeding, such as paying attention to the appropriate dose of antithrombotic drugs.

The high Killip classification is associated with an increased risk of bleeding.^[3,10, 11]^ In our study, patients with Killip class IV were at 5.1 higher risk of in-hospital bleeding than patients with Killip class I~III. Insufficient tissue perfusion adversely affects the coagulation system and platelet function.^[11]^ Insufficient tissue perfusion may cause gastritis or ulceration and increase the possibility of gastrointestinal bleeding.^[11]^

Advanced age has been reported to be an independent risk factor of bleeding.^[3, 11–15]^Age may change the balance between the risks and benefits of treatment strategies.^[16]^ The cause of the higher risk of bleeding in the elderly may be multifactorial, including decreased kidney function and increased sensitivity to anticoagulants.^[17]^ It is speculated that the presence of local vascular changes is a potential explanation for the increased incidence of bleeding complications in elderly patients.^[6]^ It is recommended that elderly patients have stomach protection.^[18]^

Moscucci et al. observed that seniors, women, history of bleeding and renal insufficiency were independent predictors of major bleeding among the patients with 8151 STEMI, 7440 non-ST elevation myocardial infarction (NSTEMI) and 8454 unstable angina registered in the Global Acute Coronary Events Registry (GRACE).^[6]^ Spencer et al. found that major bleeding occurred in 2.8% of 40, 087 patients with AMI enrolled in the GRACE. These patients were older, more severely ill, and more likely to undergo invasive procedures.^[19]^ The Can Rapid Risk Stratification of Unstable Angina Patients Suppress Adverse Outcomes With Early Implementation of the ACC/AHA Guidelines (CRUSADE) bleeding score was used to stratify the risk of major bleeding in NSTEMI patients; Subherwal et al. used 71,277 patients to derive and 17,857patients to validate a model that identifies 8 independent baseline predictors.^[20]^ Nikolsky et al. found age >55 years, female gender, estimated glomerular filtration rate < 60 mL/min/1.73 m^2^, pre-existing anaemia, administration of low-molecular-weight heparin within 48 hour pre-PCI, use of glycoprotein IIb/IIIa inhibitors, and intraaortic balloon pump use were independent predictors of bleeding.^[17]^

Our diagnostic model of in-hospital bleeding builds upon these studies in several ways. It is not a relative value but an absolute value. It includes only baseline factors, including age and Killip classification. It is easily calculated at patient presentation. It can retain discriminatory, thereby improving its effectiveness in clinical decision-making no matter what treatment is used (such as invasive care or antithrombotic drugs). It was developed in unselected real-world populations, including those who received initial invasive strategies and revascularization, and those who were conservatively treated without catheterization. Algorithms that can help doctors evaluate the diagnosis should be simple and easy to apply to the bedside, and should use clinical data routinely provided by the hospital.The nomogram we constructed for in-hospital bleeding captures most of the diagnostic information provided by the complete logistic regression model and is easier to use at the bedside.

### Study Limitations

This is a single center experience. Some patients were selected >10 years ago, so their treatment may not meet current standards and techniques.It does not include bleeding related to catheterization. The use of antithrombotic drugs and previous bleeding history were not obtained in this study, so we cannot determine the effect of anticoagulation and previous bleeding history on bleeding risk.Finally, the c statistic of the study in-hospital bleeding model at 0.777 in the derivation and 0.7234 in the validation cohort is modest.

## Conclusions

We developed and externally validated a moderate diagnostic model of in-hospital bleeding in patients with acute STEMI.

### Abbreviations

AMI = acute myocardial infarction; AF = atrial fibrillation;AIC = Akanke information criterion;AUC = area under the receiver operating characteristic curve; AVB = atrioventricular block; BIC = Bayesian information criterion; CABG = coronary artery bypass grafting; CI = confidence interval; CKD = chronic kidney disease; CRUSADE = Can Rapid Risk Stratification of Unstable Angina Patients Suppress Adverse Outcomes With Early Implementation of the ACC/AHA Guidelines;HCD = history of cerebrovascular disease; GRACE = global registry of acute coronary events registry; MI = myocardial infarction; NSTEMI = non ST elevation myocardial infarction; PCI = percutaneous coronary intervention; ROC = receiver operating characteristic;STEMI = ST elevation myocardial infarction; TIMI = Thrombolysis in Myocardial Infarction;TRIPOD = Transparent Reporting of a multivariable prediction model for Individual Prognosis Or Diagnosis.

## Data Availability

All data generated or analysed during this study are included in this published article [and its supplementary information files].

Supplementary materials

The data are demographic, and clinical characteristics of hospitalized patients with acute STEMI. AGE=age; ALLAF=atrial fibrillation; AVB=atrioventricular block; BLOOD=all-cause bleeding; CABG =history of coronary artery bypass grafting; CKD=history of chronic kidney disease; DM=history of diabetes; HBP = history of hypertension; HCD= history of cerebrovascular disease; HPCI=history of percutaneous coronary intervention; KI = Killip I; KII = Killip II;KIII = Killip III; KIV = Killip IV; OMI=history of myocardial infarction; PCI=underwent PCI during hospitalization; S = sex.

https://pan.baidu.com/s/1IUfgPFqZbPhV__AHd5oqVA

## Acknowledgments

Not applicable.

## Declarations

### Ethical approval and consent to participate

Ethic committee approved the study. Name of the ethic committee:Ethics committee of Beijing Anzhen Hospital Capital Medical University. Approved No. of ethic committee: 2019044X. It was a retrospective analysis and informed consent was waived by Ethics Committee of Beijing Anzhen Hospital Capital Medical University.

### Statement of human and animal rights

All procedures performed in studies involving human participants were in accordance with the ethical standards of the institutional and/or national research committee and with the 1964 Helsinki declaration and its later amendments or comparable ethical standards. The study was not conducted with animals.

### Consent to publish

Not applicable.

### Availability of data and materials

#### Supplementary materials

The data are demographic, and clinical characteristics of hospitalized patients with acute STEMI. AGE = age; ALLAF = atrial fibrillation; AVB = atrioventricular block; BLOOD = all-cause bleeding; CABG = history of coronary artery bypass grafting; CKD = history of chronic kidney disease; DM = history of diabetes; HBP = history of hypertension; HCD = history of cerebrovascular disease; HPCI = history of percutaneous coronary intervention; KI = Killip I; KII = Killip II;KIII = Killip III; KIV = Killip IV; OMI = history of myocardial infarction; PCI = underwent PCI during hospitalization; S = sex.

### Authors’ contributions

Yong Li contributed to generating the study data, analysed, interpreted the study data, drafted the manuscript, and revised the manuscript. Yong Li is being responsible for the overall content as guarantor. All authors read and approved the final manuscript.

